# In-Patient Repeatability and Sensitivity Study of Multi-Plane Super-Resolution Ultrasound in Breast Cancer

**DOI:** 10.1101/2024.10.15.24315514

**Authors:** Megan Morris, Emily Durie, Victoria Sinnett, Matthieu Toulemonde, Ioannis Roxanis, Steven Allen, Kate Downey, Julie Scudder, Tanja Gagliardi, Pauline Scott-Mackie, Samantha Nimalasena, Jipeng Yan, Biao Huang, Joseph Hansen-Shearer, Lone Gothard, Justine Hughes, Matthew D Blackledge, Navita Somaiah, Meng-Xing Tang

**Author notes:** Joint senior and corresponding authors: Meng-Xing Tang -, Navita Somaiah – and Matthew D Blackledge –.

## Abstract

**Purpose:** Super-resolution ultrasound (SRUS) is a promising imaging modality for detecting early microvascular changes after cancer treatment, offering advantages over tumour-size methods to evaluate response. For clinical application, it is crucial to assess repeatability of SRUS-derived biomarkers and their sensitivity to post-treatment changes.

**Experimental Design:** Clinical data were collected from breast cancer patients undergoing radiotherapy. 24 repeatability scans were conducted, and 11 participants underwent SRUS response assessment at 2-weeks and 6-months post-radiotherapy. Ultrafast CEUS acquisitions sampled four imaging planes of each tumour, generating 2D SRUS maps of microvascular structure and dynamics. SRUS-derived quantitative parameters were extracted, with repeatability assessed using the Repeatability Coefficient (RC). Changes in quantitative parameters were analysed post-radiotherapy, and the RC defined significant changes. SRUS-derived quantitative parameters were compared to histopathological CD31 staining of biopsy samples.

**Results:** The RCs of SRUS quantitative parameters improved when averaged over more imaging planes, indicating improved repeatability. Significant changes in SRUS quantitative parameters were observed at 2-weeks post-RT in 5/11 participants. In contrast, only 1/11 participants showed significant tumour size changes. By 2-weeks or 6-months post-RT, significant changes in SRUS quantitative parameter were detected in all participants, while significant changes in tumour size were observed in 6/11 participants. Among 10 participants with corresponding CD31 vessel counts, 7 showed a correlation between the direction of change in histopathological vessel count scores and SRUS vessel density.

**Conclusions:** This repeatability and response assessment study establishes multi-plane SRUS as a robust and sensitive tool for detecting early tumour microvascular changes in patients undergoing treatment.

**Funding:** CRUK Convergence Science Centre, Kortuc Inc., NHS, NIHR, ICiC, IAA.

## Introduction

Breast cancer is a global health challenge, with 2.3 million new cases annually. Most diagnoses are early, however, about 20% of cases in high-income and 50-80% in low-to middle-income countries are diagnosed at late clinical stages (Ginsburg 2017, Fuentes 2024, Coles 2024). Patients presenting in locally advanced stages require primary systemic therapy or radiotherapy for tumour control.

Monitoring treatment response is an integral part of ongoing patient care. Current standards include RECIST 1.1. (Eisenhauer 2009), which measures tumour size changes using contrast-enhanced CT and MRI scans, tracking the tumour’s longest diameter over time. Despite standardising tumour measurement, RECIST faces criticism for inter-observer variability and not fully representing tumour morphology (Gerwing 2019). Additionally, size measurements can underestimate response by failing to capture biological heterogeneity, and early micro-environmental changes occurring before changes in tumour size (Ko 2021). This is especially problematic with radiotherapy-treated breast tumours, as radiotherapy causes tissue thickening and scarring, resembling residual enhancing masses on standard imaging (Padhani 2003, Lee 2011, EL-Adalany 2016, Sarsenov 2017). Therefore, more sensitive imaging tools are needed to detect response, resistance or relapse better, thereby improving patient outcomes.

The tumour microvasculature is a promising imaging target. Solid tumours have structurally and functionally abnormal micro-vessel networks due to pathological angiogenesis (Ehling 2014). The chaotic and disorganised structure leads to regionalised chronic hypoxia, which is associated with poor outcomes due to therapy resistance (Walsh 2014, Barsoum 2014). The tumour microvasculature thus has prognostic potential, and early changes can indicate therapy response (Jia 2016, Tolaney 2015) and identify non-responders (Bullitt 2006), allowing timely treatment modifications.

To address this, clinical microvasculature imaging technologies are needed. Perfusion imaging techniques using MRI, CT and PET have helped bridge the gap between tumour size and biology (Stieb 2020). However, they provide only surrogate perfusion measures due to low spatial and temporal resolution. Advancements in ultra-fast plane-wave Doppler ultrasound imaging has improved sensitivity to smaller, slow-flow blood vessels (Demené 2015), however, the smallest micro-vessels are difficult to detect (Song 2023). High spatial resolution imaging, such as microscopy, often requires biopsies or has limited imaging depth. The ideal in vivo vascular imaging tool would surpass these limitations, offering high spatial and temporal resolution, a large field of view, and be non-invasive, non-ionising and non-toxic.

Super-Resolution UltraSound (SRUS) is a cutting-edge clinical imaging technology, with microvascular resolution (Christensen-Jeffries 2020) down to tens of micrometres while maintaining a large field of view. SRUS through localising and tracking microbubble contrast agents, also known as Ultrasound Localization Microscopy (ULM), extends the capabilities of contrast-enhanced ultrasound, a clinically established technique involving intravenous administration of microbubble contrast agents. These microbubbles illuminate the microvasculature, revealing even the smallest micro-vessels with slow blood flow velocities that non-contrast ultrasound techniques cannot detect. By combining contrast-enhanced ultrasound with post-processing algorithms, SRUS overcomes the spatial resolution limitations inherent in traditional ultrasound systems. The microvasculature is reconstructed with microscopic precision and critical features such as structure and blood flow velocities can be quantified. Since the initial demonstration of super-localisation (Siepmann 2011, Couture 2011), and super-resolved flow velocity mapping (Christensen-Jeffries 2015, Errico 2015), SRUS has garnered interest as a powerful tool to investigate diseases such as cancer in both pre-clinical (Lin 2017, Zhu 2019, Lowerison 2020) and clinical settings (Opacic 2018, Dencks 2019, Kanoulas 2019, Huang 2021, Zhu 2022).

For clinical use, SRUS-derived quantitative parameters must be robust and accurately characterise the tumour microvasculature. To the best of our knowledge, the repeatability of SRUS has not been well established in the literature. Yet, evaluating the test-retest repeatability is vital for clinical translation (O’Connor 2017). This study aimed to assess the test-retest repeatability of SRUS in breast cancer patients undergoing radiotherapy. Additionally, it aimed to identify the SRUS-derived quantitative parameters that are sensitive in detecting significant post-treatment microvascular changes. By doing so, this study contributes towards establishing SRUS as a reliable clinical assessment tool, offering a low-cost solution for monitoring patients undergoing cancer treatment.

## Methods

### Participant Population

#### Clinical Trial

The SRUS repeatability and response assessment data were generated from the randomised KORTUC Phase 2 clinical trial (ClinicalTrials.gov: NCT03946202). The KORTUC trial recruits patients with inoperable, locally advanced breast cancer. The trial aims to test the efficacy of an intra-tumoural KORTUC injection, a diluted hydrogen peroxide-based radiosensitiser, combined with radiotherapy (Nimalasena 2020). All participants received 36Gy of radiation delivered in twice weekly doses of 6Gy each, over 3 weeks, with or without KORTUC. Participants gave written consent to participate in the SRUS sub-studies and underwent SRUS imaging pre-radiotherapy (RT), and at 2-weeks, 6-, 12-, 18- and 24-months post-RT to assess treatment response. Post-RT refers to the end of the entire radiotherapy treatment period (3 weeks). Ethical approval was granted by the West of Scotland Research Ethics Committee (REC reference 20/WS/0019).

#### SRUS Repeatability Study

Twelve KORTUC trial participants were recruited between February 2022 and March 2024, with 24 repeatability scans conducted. Eight participants had separate repeatability scans at multiple time-points. In addition to the typical practice of pre-treatment repeatability studies (O’Connor 2017), we also wanted to ensure SRUS repeatability post-treatment. Five participants had repeatability scans at baseline, eight at 2-weeks, one at 3-months, four at 6-months, and two at 12-, 18- and 24-months post-RT. The repeatability scans from the same participant at different time-points were considered independent during statistical analysis, due to the post-treatment changes that occurred over that time. Repeatability was assumed to be consistent across all imaging time-points. A repeatability scan was completed in one 20-minute imaging session.

#### SRUS Response Assessment Study

Eleven KORTUC trial participants were recruited between December 2021 and May 2024. SRUS imaging was performed at baseline, 2-weeks, and 6-months post-RT for response assessment. Further time-points post-RT were not included in this study as the focus was on investigating early microvascular changes. The participant population for the SRUS repeatability and response assessment studies is summarised in Figure 1.

**Figure 1.**
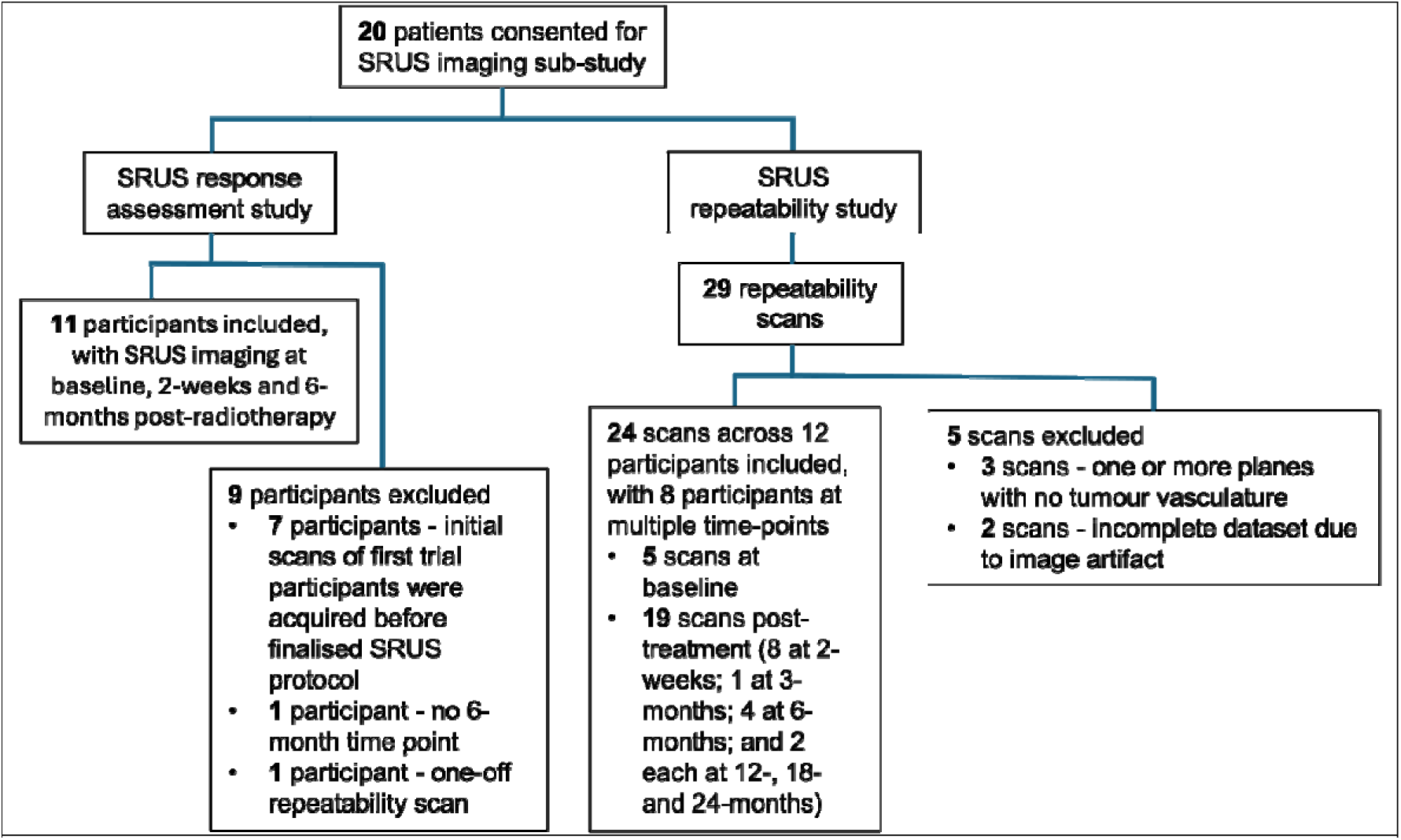
The participant population for the SRUS imaging sub-studies, detailing those who were included in the SRUS repeatability study and response assessment study, and reasons for exclusion.

### Ultrasound Equipment and Acquisition Settings

Ultrasound acquisitions used a GE L3-12-D linear array transducer (GE HealthCare Technologies Inc., Chicago, IL, USA) and a Verasonics Vantage 256 research scanner (Verasonics Inc., Kirkland, WA, USA). Amplitude-modulated plane-waves were transmitted at 5MHz with -10 to 10° steering angles, with a mechanical index of 0·1. Data was captured at compounded frame-rates of 100 frames per second (fps) for 5s of acquisition. The number of compounded angles varied between but not within participants, from 4-10. Ten compounded angles were mostly used, to create a high-quality B-mode image for tumour segmentation (20/24 repeatability scans in the repeatability study; 8 out of 11 participants in the response assessment study).

Hardware constraints led to a trade-off between compounded frame-rate, length of acquisition, and number of compounded angles, due to a limit on data collection per acquisition. Three participants within the response assessment study and four scans within the repeatability study had a reduced number of compounded angles (4-8) to achieve higher compounded frame-rates (200-400 fps). These frame-rates were reduced to 100 fps during post-processing so as not to impact the repeatability statistics and response assessment.

### Imaging Protocol

The imaging protocol is illustrated in Figure 2c. A 2·5mL SonoVue bolus was injected intravenously over 15s followed by a saline flush. Ultrasound acquisitions began 35s post-injection. The probe was hand-held horizontally by a consultant breast radiographer at the centre of the tumour, then moved to a 90°, 45° clockwise and 45° counterclockwise imaging plane with the tumour centre re-located. For the repeatability scan, the imaging protocol was repeated 5 minutes after the initial bolus injection.

**Figure 2.**
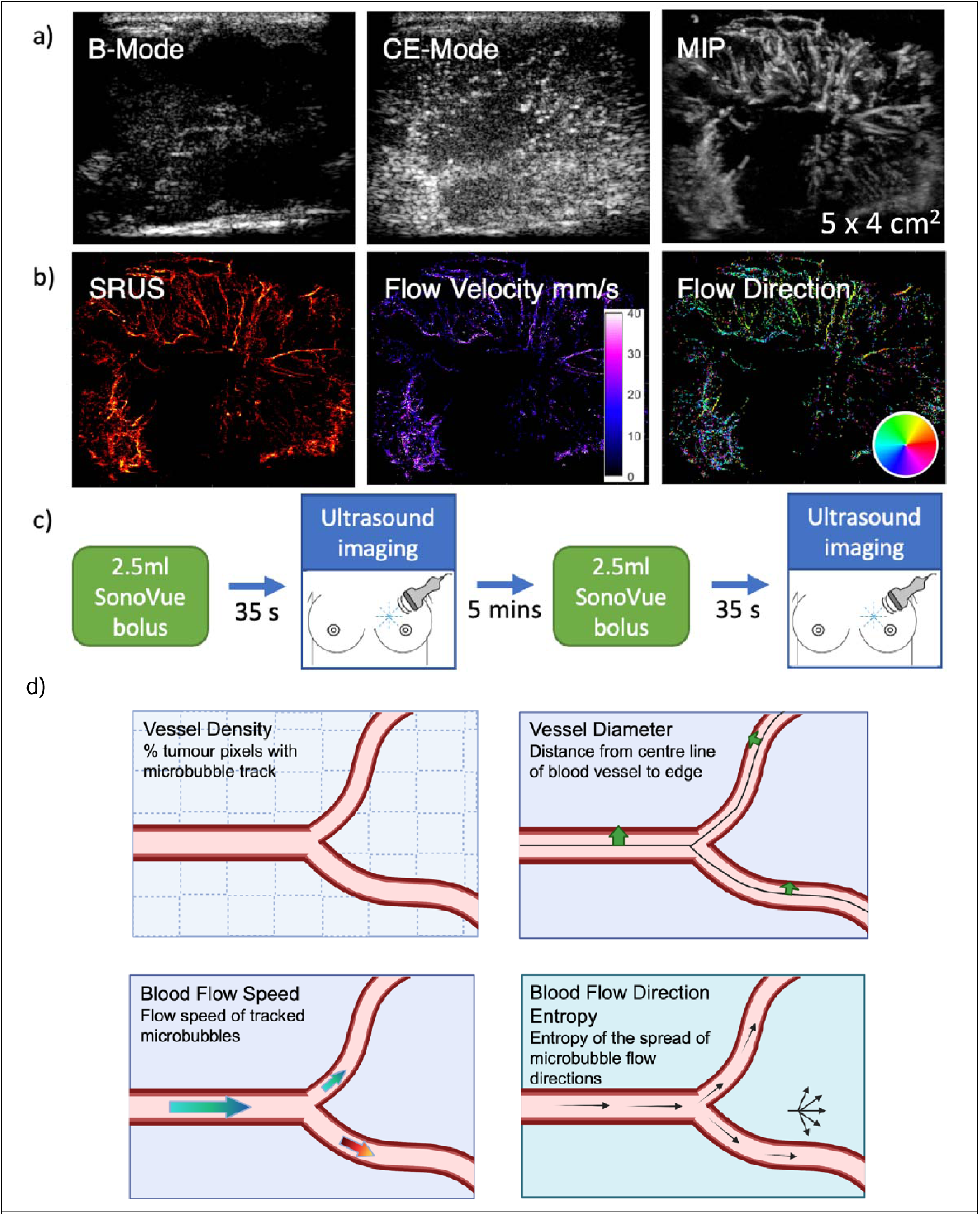
**a)** The current clinical capabilities of ultrasound with intravenous contrast for breast tumour imaging; including, a B-mode tissue image, the corresponding contrast-enhanced image (single frame), and the maximum intensity projection (MIP) of the processed contrast-enhanced data. **b)** The corresponding super-resolution ultrasound maps, reconstructed from the contrast-enhanced ultrasound scan. Contrast tracking provides blood flow speed and direction maps. **c)** The super-resolution ultrasound imaging protocol. A 2.5mL bolus of SonoVue contrast was intravenously injected. After 35 seconds, ultrasound acquisitions were acquired in the horizontal plane, then at 90°, 45° clockwise and 45° counterclockwise at the tumour centre. For the repeatability scan, the imaging protocol was repeated 5 minutes after the initial bolus injection. **d)** Diagramatic depiction of the SRUS quantitative parameters.

When designing the imaging protocol, a 2.5mL bolus gave sufficient microbubbles to reconstruct the microvasculature, along with isolated microbubbles for SRUS localisation and tracking. Based on visual inspection of the real-time feedback, the microbubble bolus reached peak concentration at around 35s, which is when the first acquisition was taken. Despite differences in microbubble concentration between the first and last imaging plane, the order of the imaging planes remained consistent. Each plane was therefore comparable to that same plane in repeat scans or in scans at different imaging time-points.

Participants breathed normally during the scan, and the 5-second acquisition time was short enough for the radiographer to hold the probe still with minimal out-of-plane motion. The stiffness of tumours and post-therapy fibrotic tissue meant that the data was minimally affected by breathing motion. Any data with significant motion artefacts were excluded, which only affected 2 repeatability scans (Figure 1). Before injecting the second bolus, the clearance of the first bolus was visually confirmed on the real-time feedback. A 5 minute-interval between bolus injections allowed for the microvasculature to clear.

Each SRUS imaging session lasted around 20 minutes. Set-up took around 10 minutes, including cannulation, a preliminary clinical ultrasound, and SonoVue preparation. The clinical ultrasound was required to locate the tumour, due to better real-time B-mode image quality. The clinical probe was replaced by the research probe, and the tumour centre was re-located. The SRUS imaging from the start of the microbubble injection to the end of the multi-plane imaging took under 2 minutes, which is approximately the duration of usable contrast. For a repeat scan, a second bolus injection was administered with a 5-minute interval between injections. The total imaging time was under 10 minutes. When transitioning between imaging planes, the radiographer only required about 5s to find the central slice of the tumour using the B-mode feedback.

### Contrast-Enhanced Data Processing

The raw ultrasound data was reconstructed with a 100×30·8µm^2^ pixel size in the lateral and axial directions, respectively. An example of a B-mode image of a tumour is shown in Figure 2a, alongside a single contrast-enhanced frame before data processing. In-plane motion correction was applied to minimise the effects of the hand-held probe and breathing motion. A non-rigid registration algorithm estimated and applied motion correction from B-mode images to contrast-enhanced (CE-) data (Rueckert 2009).

A moving average was subtracted to remove residual tissue signal. The optimal window size (∼0·11s) was determined with consideration of the minimum blood flow speed and residual tissue motion. By visually inspecting the maximum intensity map of the processed data, smaller window sizes removed slow-moving microbubbles, and higher window sizes did not remove all residual tissue, and regardless, no further blood vessels were revealed. Based on the frequency response of the moving average filter, the half-power (-3 dB) cut-off frequency is 6.9 Hz. Using the Doppler frequency formula, and given an acquisition frame rate of 100 fps and transmission frequency of 5 MHz, the axial cut-off velocity for this window size is 1 mm/s. The same window size was used across the datasets.

Then, after envelope detection, the data was spatially (300µm width) and temporally (3 frames) smoothed using a 3D Gaussian filter (imgaussfilt3, MATLAB, Image Processing Toolbox) to improve microbubble visualisation and reduce noise, followed by log-compression. The noise background was estimated from an air acquisition, with the same acquisition settings and processed the same as above. The maximum intensity projection of the processed air acquisition gave the upper noise threshold. This threshold was subtracted from the clinical data, with values below zero considered as noise. An example of a maximum intensity projection of processed contrast-enhanced data is shown in Figure 2a.

### SRUS Processing

SRUS maps were reconstructed with a 3·1µm lateral/axial resolution grid. Microbubble properties (area, power, eccentricity, unimodality and solidity) were specified, to differentiate microbubbles from remaining clutter. The microbubble property limits were chosen by manual selection of microbubbles across different participants and scans, to identify the range of values.

The SRUS algorithm localised isolated microbubble centres and tracked them between frames using a Kalman motion model-based tracking algorithm (Yan 2022, Yan 2023). Accumulation of localisations and tracks over time generated super-resolved maps of microvascular structure, blood flow speed and direction, with an example shown in Figure 2b. All acquisition frames were used for SRUS reconstruction, as there was minimal out-of-plane motion across the dataset. Any data with significant motion artefacts were excluded, however, this only affected 2 repeatability scans (Figure 1). Significant motion artefacts were identified by tissue signal breakthrough after the moving average subtraction.

A Gaussian filter (imgaussfilt, MATLAB, Image Processing Toolbox) was applied with a standard deviation related to the localisation uncertainty, calculated at 20μm. The localisation uncertainty was determined by imaging a stationary wire in a water tank over 100 frames, using the same acquisition settings as discussed above. The stationary wire, which simulates a microbubble scatterer, was localised across the frames. The wire was imaged at various positions within the imaging field, and the standard deviation of the x and y localisation coordinates was calculated for these positions. The average standard deviation was then calculated across all positions, and was larger along the x-axis, with a value of 20μm, which was taken as the localisation uncertainty. This value of 20μm was consistent regardless of the number of compounded angles.

### Quantitative Parameters

Tumours were manually delineated using the B-mode tissue images (Figure 2a) and verified by a consultant breast radiographer. The SRUS quantitative parameters were extracted from the super-resolved localisation, blood flow speed and blood flow direction maps, within the tumour region. The quantitative parameters are described below and depicted in Figure 2d.

#### Vessel Density

Vessel density was calculated as the percentage of pixels within the delineated tumour that a microbubble has passed through, determined by microbubble localisations and tracks. Tumours have denser vessel areas than healthy tissue due to neo-angiogenesis.

#### Vessel Diameter

Vessel diameter was calculated by first skeletonising the binary image of the microvascular structure (bwmorph, MATLAB, Image Processing Toolbox) to obtain the blood vessel centrelines. The distance from each pixel on the centreline to the nearest edge of the blood vessel was computed (bwdist, MATLAB, Image Processing Toolbox), to determine the blood vessel’s radius at each point. The vessel diameter at each point was then calculated as twice the radius. The in-plane mean was calculated by averaging the diameters over all points along the vessel’s centreline. Tumours have larger microvessel diameters than healthy tissue (Nagy 2009, Forster 2017).

#### Blood Flow Speed

The blood flow speed values were extracted from the super-resolved maps of the blood flow speed obtained from microbubble tracking. Blood flow speed was calculated as the distance between linked microbubbles divided by the frame-rate. The in-plane mean was calculated.

#### Local Blood Flow Direction Entropy

The local flow direction entropy quantifies the organisation of the microvasculature (Opacic 2018). Local blood flow direction entropy was calculated using the super-resolved maps of the blood flow direction from microbubble tracking, by sub-dividing the tumour into smaller areas of 1.6×1.6mm, which is on the scale of micro-metastases. Blood flow direction is the angle (from 0 to 360°) between linked microbubbles. The blood flow directions are binned into 180 bins covering 0 to 360°, and entropy was calculated as 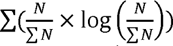, where N is the bin count. Tumour blood vessels can be tortuous and branch chaotically (higher entropy), compared to the linearity and order in healthy tissue (lower entropy).

#### CEUS Sum of Intensities

A contrast-enhanced ultrasound quantitative parameter was extracted from the CEUS processed data, specifically, the sum of intensities within the maximum intensity projection divided by the tumour area, for comparison with the SRUS quantitative parameters.

#### RECIST Measurement

The percentage change in the longest tumour diameter was measured by radiologists according to RECIST 1.1. criteria, on MRI scans taken at the same time-point as the SRUS imaging. A percentage decrease greater than 30% is considered a partial response, and a percentage increase greater than 20% is considered as progressive disease. In between this is stable disease.

Fractal dimension, distance to vessel and distance to perimeter were also evaluated but were not sensitive to treatment response. These parameters have been described in the supplementary methods.

### Biopsies and Immunohistochemical Staining

Duplicate core biopsies were obtained from 10 participants at baseline and 2-weeks post-RT, from two non-necrotic areas of the tumour to capture heterogeneity, guided by clinical ultrasound. Biopsies were processed into formalin fixed paraffin embedded blocks (FFPE). Immunohistochemical staining for endothelial cell marker CD31 (clone JC70A) was performed on 3µm sections using a Leica Bond III platform and Bond Refine Detection Kit (HSL Laboratories). Slides were digitised at 40X magnification using a Nanozoomer© (Hamamatsu) digital slide scanner. A consultant breast histopathologist (IR) scored CD31 staining, with 1=0-5; 2=5-10; 3=10-15; 4=15-20; 5=20-25; 6=25-30; 7=30-35 vessels per 0·5mm^2^. Mean scores were calculated from duplicate biopsies.

### Statistical Analysis

For the SRUS repeatability study, the repeatability coefficient (RC) was calculated for each SRUS quantitative parameter, *x*_i}_, where *x_ij_*;= 1,…,*N* represents the repeatability scan (N=24) and *<u>j</u>* = 1,2 is the test-For the SRUS repeatability study, the repeatability coefficient (RC) was calculated for each SRUS retest measurement, i.e., the quantitative parameter after the first and second bolus. The natural logarithm of the quantitative parameters was used for statistical analysis, as initial analysis revealed heteroskedastic repeatability, i.e., the variability was related to the mean, with more vascular tumours exhibiting relatively poorer absolute repeatability. The repeatability coefficient was calculated as:

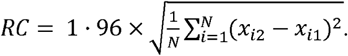

Tumours provided four measurements per quantitative parameter due to multiple imaging planes acquired during the scan. The repeatability coefficient was calculated for all the imaging plane combinations, to investigate the impact of averaging across the imaging planes on repeatability. In other words, quantitative parameter repeatability was investigated for a single imaging plane, and then for a combination of two, three and four imaging planes.

For the SRUS response assessment study, the changes in the quantitative parameters (averaged across the four imaging planes) at 2-weeks and 6-months post-RT were analysed to assess their potential as sensitive response biomarkers. The repeatability coefficient was used to evaluate significant change. The analysis was conducted blind to the treatment group allocation and clinical outcome as the main clinical trial is still ongoing.

## Results

### Super-Resolution Ultrasound Demonstrated Repeatable Detection of Microvascular Structures Across Multiple Imaging Planes

From the SRUS repeatability study, the super-resolved maps acquired after the first and second boluses revealed matching microvascular structures, as illustrated in Figure 3 for an example participant. This consistency was evident across multiple imaging planes of the tumour (0° (horizontal), 90°, 45° clockwise and 45° counterclockwise). The white dashed boxes in the super-resolved maps highlight these consistently located structures, demonstrating the capability of an experienced breast radiographer to repeatedly pinpoint the tumour centre across multiple imaging planes.

**Figure 3.**
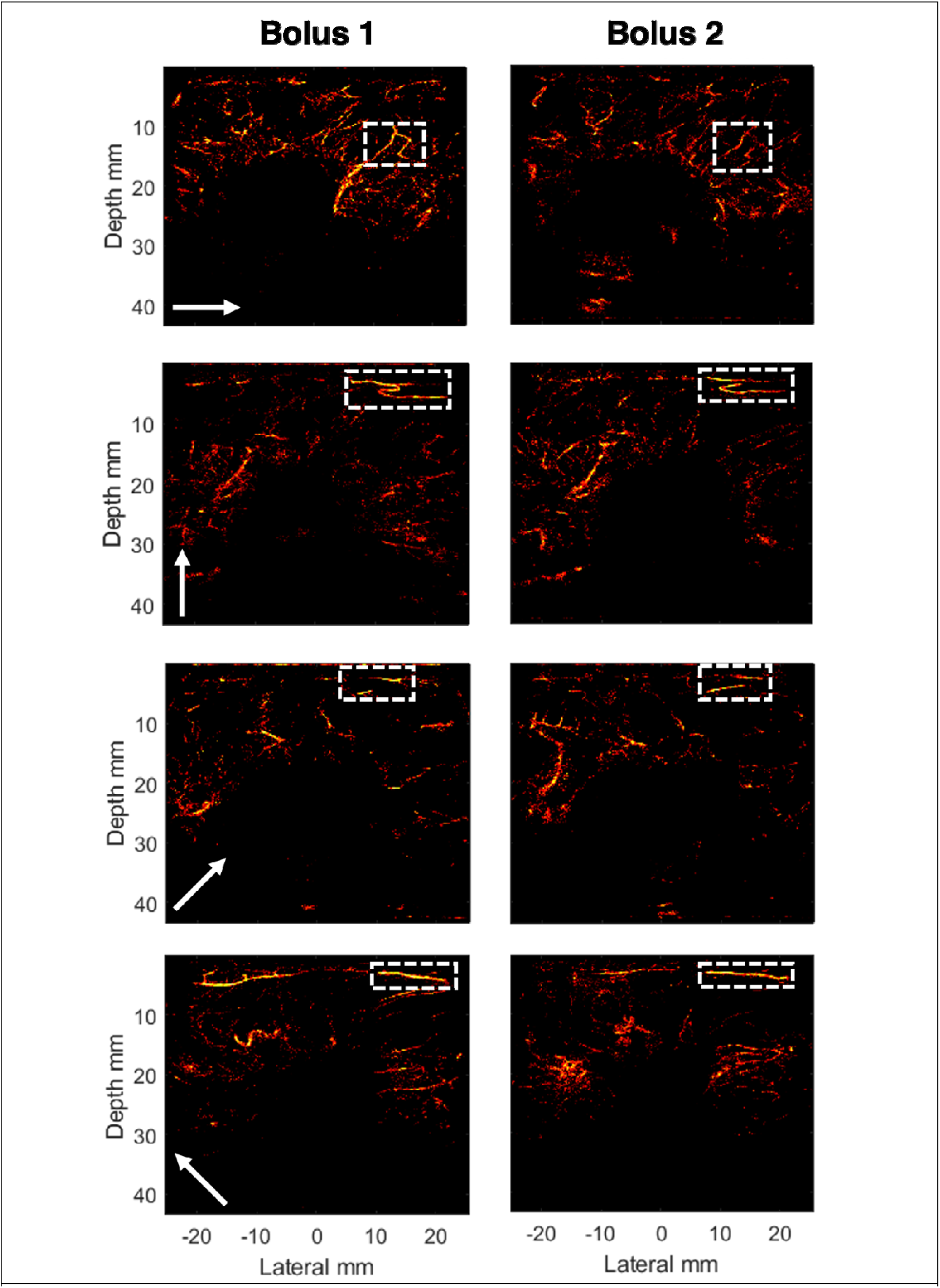
Example of a super-resolution ultrasound repeatability scan. Super-resolved maps of the breast tumour microvasculature are acquired at 4 imaging planes (horizontal, 90o, 45o clockwise and 45o counterclockwise at the tumour centre), repeated twice after a first and second contrast bolus. The brightness corresponds to the number of localisations per pixel. The white arrows show the positioning of the ultrasound probe. The white boxes highlight distinctive microvascular structures which are present after both boluses.

### Enhanced Repeatability of SRUS Quantitative Parameters Through Multi-Plane Imaging

The study examined how the repeatability coefficients (RCs) of the SRUS quantitative parameters were influenced by the number of imaging planes used for averaging the measure within the tumour. Figure 4 and Supplementary Figure 1 illustrates the effect of increasing the number of imaging planes on the RCs. Across all quantitative parameters, there was a consistent trend: the RCs decreased as the number of imaging planes used for averaging increased. This reduction in RCs was attributed to a decrease in within-subject variability. Thus, averaging across a larger number of imaging planes achieved more repeatable and robust quantitative parameters.

**Figure 4.**
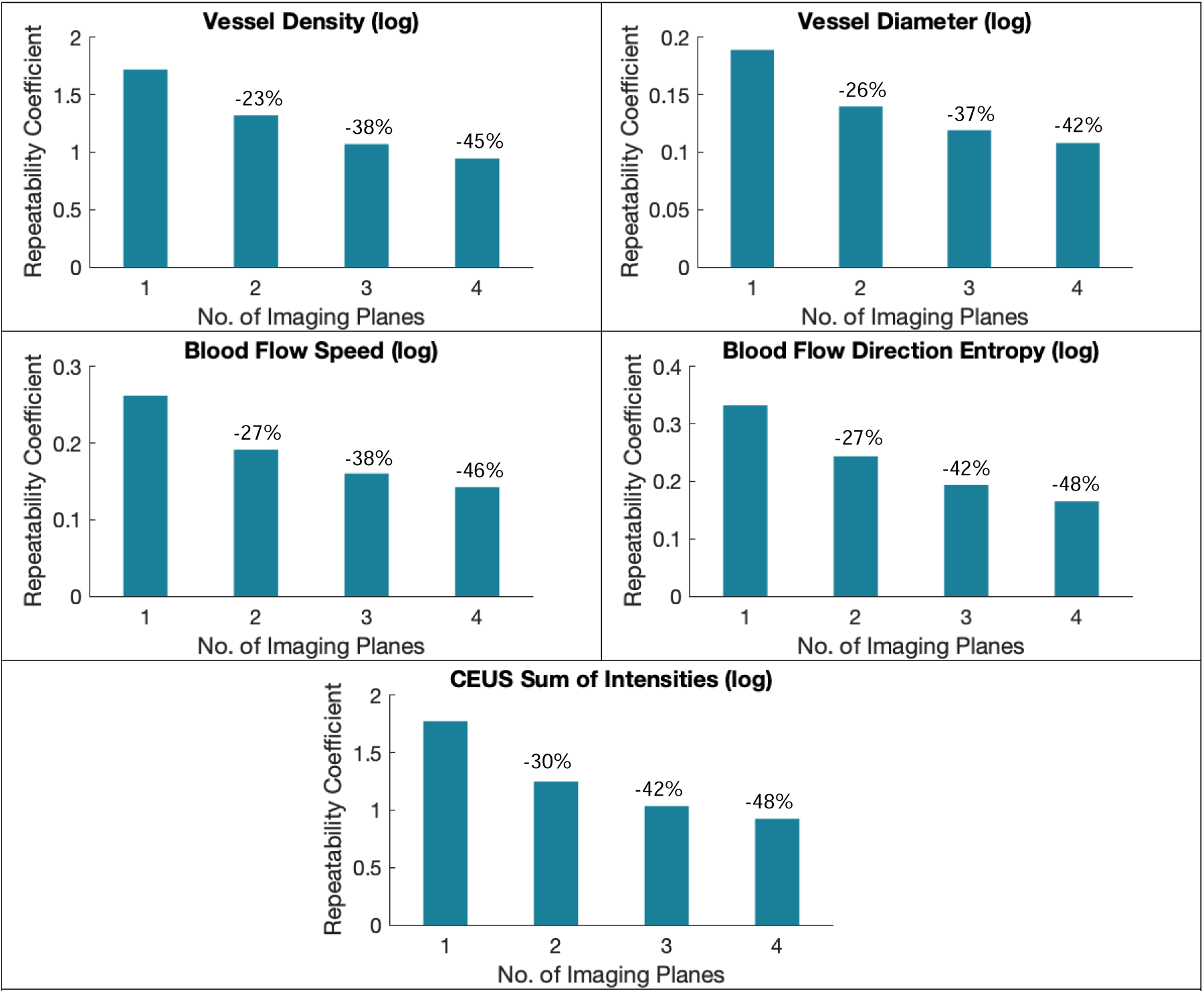
The repeatability coefficient (RC) of the natural logarithm of the SRUS quantitative parameters, as a function of the number of imaging planes used for the average measure. The percentages displayed above the bars represent the percentage change compared to using 1 imaging plane.

In comparison to measurements from a single imaging plane, when averaging over 2 imaging planes, the RCs decreased by 22-27% for the SRUS quantitative parameters (vessel density, vessel diameter, blood flow speed, local blood flow direction entropy and fractal dimension). When averaging over 3 and 4 imaging planes, the RCs decreased by 36-42% and 42-48%, respectively. Distance to vessel and distance to perimeter decreased by smaller percentages in comparison to the other quantitative parameters with increasing number of imaging planes (2: 20-24%; 3: 28-32%; 4: 32-36%). The CEUS quantitative parameter, the sum of intensities, followed a similar trend, with RCs decreasing by 30-48% when averaging over 2-4 imaging planes.

Although the RCs continued to decrease by incorporating a larger sample of imaging planes, the reduction became less pronounced. This could reflect convergence on a representative value for the entire tumour, and/or be due to the diminishing microbubble concentration over time. Nonetheless, these results underscore the advantages of a multi-plane imaging approach for achieving more reliable and consistent SRUS quantitative parameters.

### Bland-Altman Analysis Showed that SRUS Vessel Diameter and Blood Flow Speed are Highly Repeatable

Figure 5 and Supplementary Figure 2 displays Bland-Altman plots for the SRUS quantitative parameters (X), averaged over four imaging planes for 24 repeatability scans. Perfect repeatability would result in equal test-retest measurements, yielding a difference (X_2_ – X_1_) of 0. For vessel diameter, blood flow speed, fractal dimension, distance to vessel, and distance to perimeter, no apparent bias was observed, as 0 fell within the 95% confidence interval of the mean of the differences (shown by the blue shaded area). The dashed line represents the mean of the differences (). However, for vessel density and local blood flow direction entropy, 0 was just below the 95% confidence interval, indicating a potential bias. For the CEUS quantitative parameter, the sum of intensities, there was no apparent bias, though the 0 line was at the edge of the lower bound of the 95% confidence interval.

**Figure 5.**
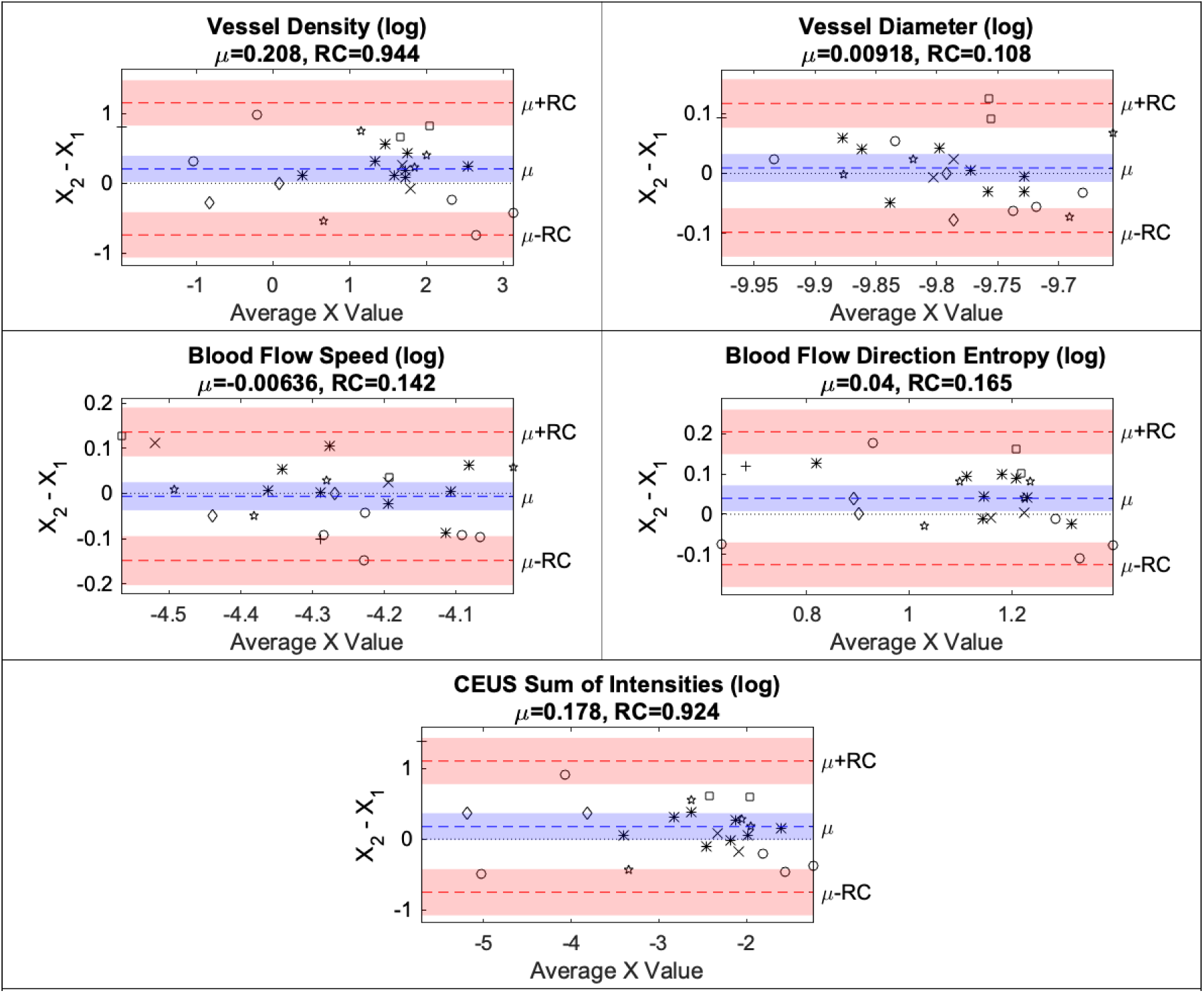
Bland-Altman plots for 24 repeatability scans. The natural logarithm of the SRUS quantitative parameters is used for statistical analysis. The y-axis is the difference between the quantitative parameter X, averaged across all 4 imaging planes, from the 2 repeat measurements (after the first bolus X_1_ and second bolus X_2_). The x-axis is the average value of the 2 repeat measurements of the quantitative parameter. The mean of the differences (μ) is given as the blue dashed line, with its 95% confidence interval being the shaded blue area. The dotted line represents 0 difference between the test-retest measurement. The mean of the differences +/-the repeatability coefficient (μ ± *RC*) is given as the red dashed line, with the 95% confidence interval of the repeatability coefficient represented by the shaded red area. The markers are the individual repeatability scans (N=24), with ○ representing baseline scans; ∗ 2-weeks post-RT; + 3-months post-RT; ⍰ 6-months post-RT; × 12-months post-RT; □ 18-months post-RT; and ⍰ 24-months post-RT.

Repeatability was assumed to be consistent for all imaging time-points, which ranged from pre-treatment to 24-months post-treatment. Within the Bland-Altman plots, different markers were used to denote different treatment phases. There is no clear evidence that repeatability varied across time-points. However, it was not statistically feasible to investigate this further due to the low sample size at each time-point. The markers were visually well mixed across the different imaging time-points for all the quantitative parameters. This was expected because baseline tumours exhibit substantial inter-patient heterogeneity.

### SRUS Quantitative Parameters Revealed Heterogeneous Tumour Microvascular Response Post-Radiotherapy

Figure 6 displays three examples of super-resolution ultrasound (SRUS) microvascular maps of the tumour and surrounding tissue, taken at baseline and 2-weeks after radiotherapy (RT). The baseline microvascular structures showed significant heterogeneity, and the post-treatment responses varied among participants. Figure 7 and Supplementary Figure 3 illustrates changes in the SRUS quantitative parameter at 2-weeks and 6-months post-RT for 11 participants, where a statistically significant change was one greater than the repeatability coefficient.

**Figure 6.**
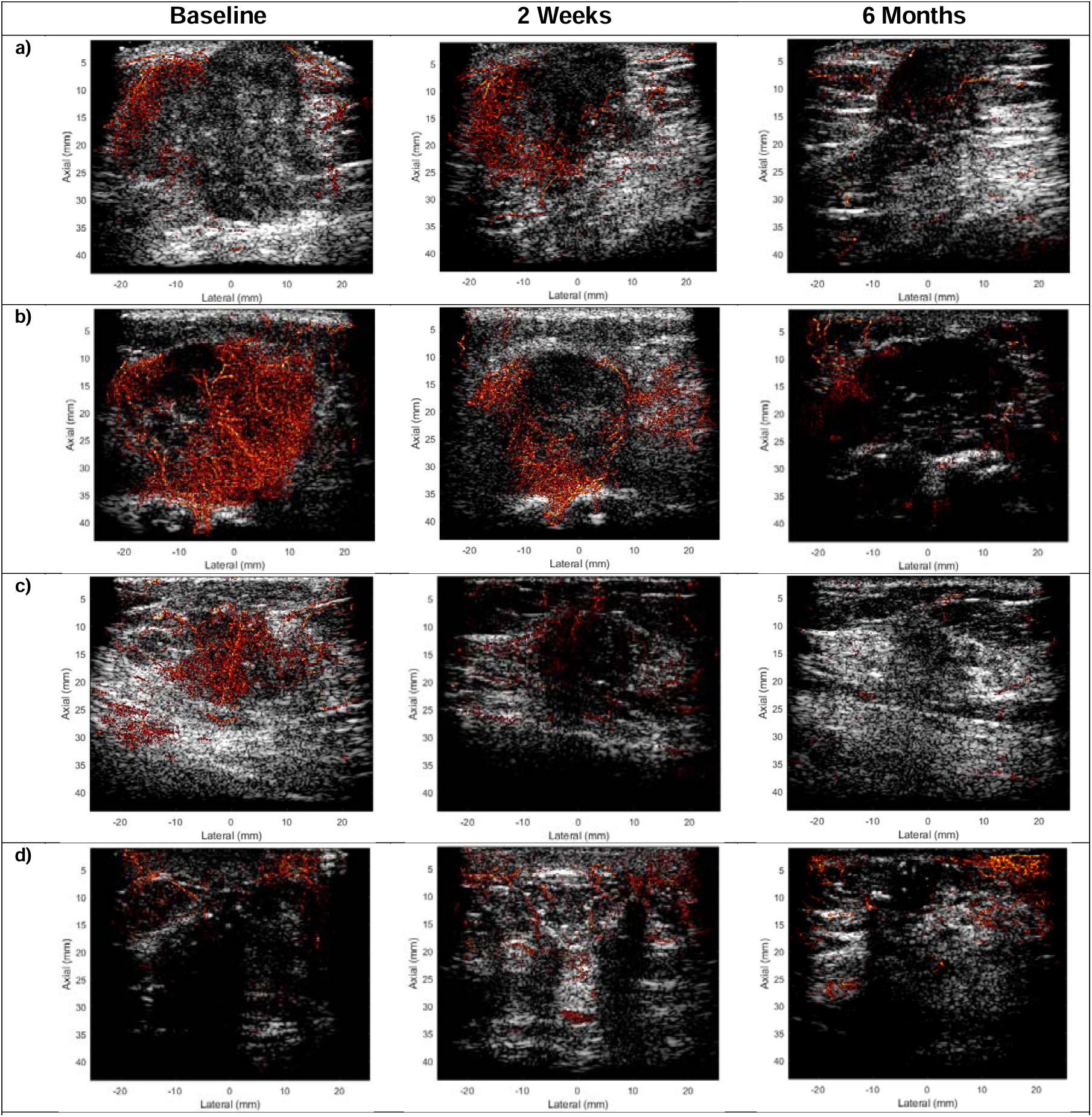
Depiction of four examples of super-resolution ultrasound microvascular maps within the tumour and surrounding tissue, overlaid on the B-mode tissue image, both before, 2-weeks and 6-months after radiotherapy (RT). The brightness corresponds to the number of localisations per pixel. Within the participant cohort, baseline tumour microvascular structures exhibit significant heterogeneity, and post-treatment responses vary widely. **a**) The baseline tumour has microvessels around the periphery but not within the core. There is a visible decrease in vascular density by 6-months post-RT. **b)** and **c**) The baseline tumour has a dense microvasculature structure. There is a visible reduction in intra-tumoural blood vessels as early as 2-weeks post-RT. **d**) The tumour exhibits a low degree of vascularity, and there is an increase in microvessels post-treatment.

**Figure 7.**
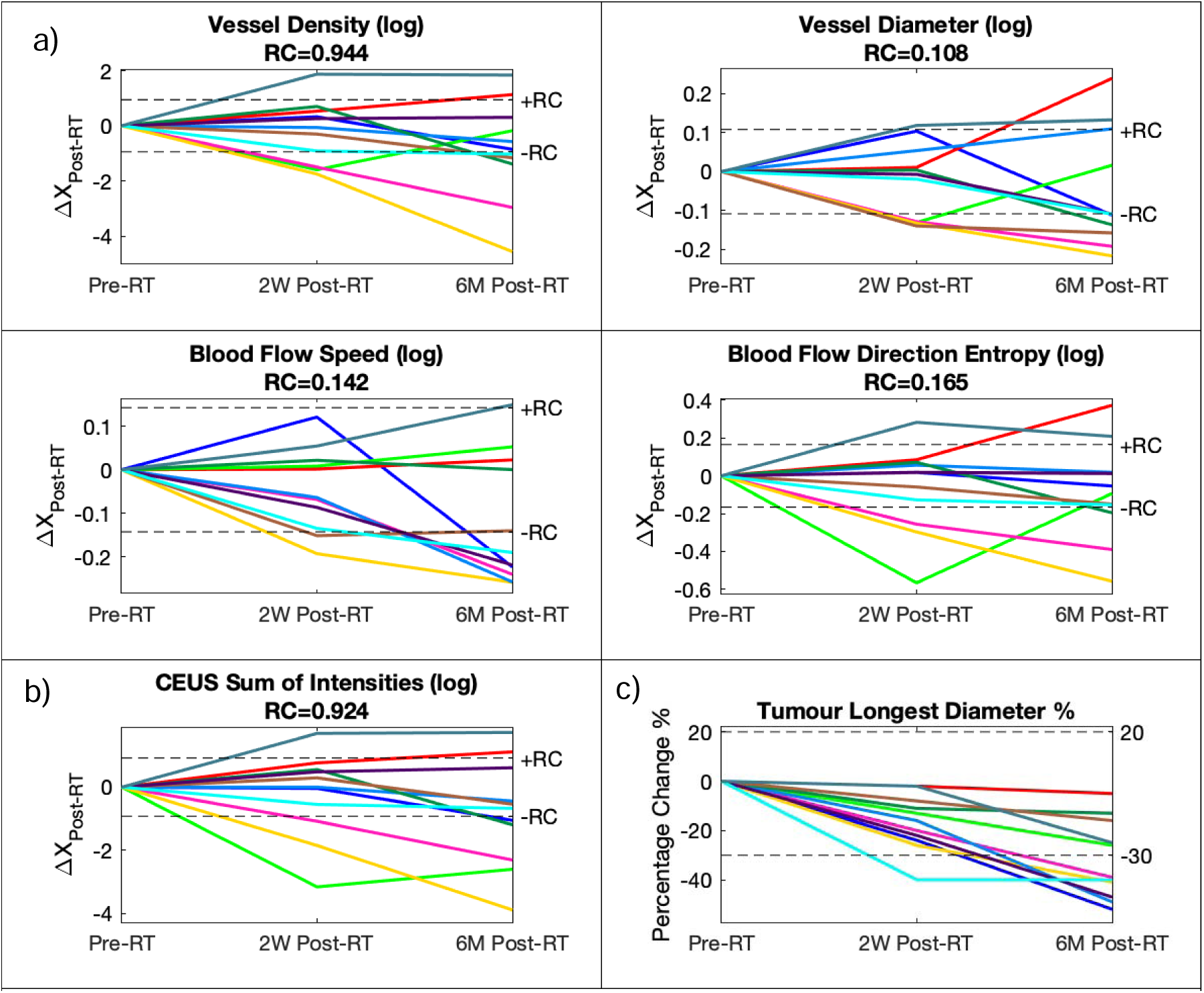
**a**) The changes of the natural logarithm of the super-resolution ultrasound (SRUS) quantitative parameters 2-weeks and 6-months post-radiotherapy (RT) from 11 participants. The y-axis shows the change in the mean of the quantitative parameter (averaged across 4 imaging planes) 2-weeks and 6-months post-RT from baseline. The dashed black lines represent the repeatability coefficient, as indicator of significant change, i.e., if the difference post-treatment is larger than the repeatability coefficient, then the change is significant. **b**) The change of the natural logarithm of a contrast-enhanced ultrasound (CEUS) quantitative parameter. **c**) The percentage change in the longest tumour diameter as measured according to RECIST 1.1. criteria, at 2-weeks and 6-months post-RT, measured by radiologists on MRI scans taken at the same time-point as the SRUS imaging. A percentage decrease greater than 30% is considered a partial response, and a percentage increase greater than 20% is considered as progressive disease. The dashed black lines represent the limits of stable disease.

#### Vessel Density

5/11 participants showed a significant decrease by 6-months post-RT, and 2 of these participants showed a significant decrease at 2-weeks post-RT. 2/11 participants showed a significant increase by 6-months post-RT, and 1 of these participants showed a significant increase at 2-weeks post-RT.

#### Vessel Diameter

7/11 participants showed a significant decrease by 6-months post-RT, and 3 of these participants showed a significant decrease by 2-weeks post-RT. 3/11 participants showed a significant increase by 6-months post-RT, and 1 of these participants showed a significant increase at 2-weeks post-RT.

#### Blood Flow Speed

7/11 participants showed a significant decrease by 6-months post-RT, and 2 of these participants showed a significant decrease by 2-weeks post-RT. 1/11 participants showed a significant increase by 6-months post-RT.

#### Local Blood Flow Direction Entropy

3/11 participants showed a significant decrease by 6-months post-RT, and 2 of these participants showed a significant decrease by 2-weeks post-RT. 2/11 participants showed a significant increase by 6-months post-RT, and 1 of these participants showed a significant increase at 2-weeks post-RT.

Across vessel density, vessel diameter and local blood flow direction entropy, one additional participant to those discussed above showed a significant decrease at 2-weeks post-RT, but had a significant increase to baseline values by 6-months post-RT. In combination, 5 of 11 participants showed a significant change at 2-weeks post-RT and all 11 participants showed a significant change by 6-months post-RT, in one or more SRUS quantitative parameters.

#### CEUS Sum of Intensities

5/11 participants showed a significant decrease by 6-months post-RT, and 3 of these participants showed a significant decrease at 2-weeks post-RT. 2/11 participants showed a significant increase by 6-months post-RT, and 1 of these participants showed a significant increase at 2-weeks post-RT.

#### RECIST Measurement

6/11 participants showed a significant decrease by 6-months post-RT, and 1 of these participants showed a significant decrease at 2-weeks post-RT. RECIST measurements showed no significant changes at 2-weeks post-RT for 3 participants who had significant decreases in 2 or more SRUS quantitative parameters and for 2 participants at 6-months post-RT. RECIST measurements showed no significant change for 2 participants who had significant increases in 3 or more SRUS quantitative parameters at either 2-weeks or 6-months post-RT. Only 1 participant had a significant RECIST measurement decrease at 2-weeks post-RT before showing a significant change in SRUS quantitative parameters at 6-months post-RT.

The direction of changes in SRUS quantitative parameters and RECIST measurements were consistent across participants. At any given time-point, participants showed either a significant decrease or no change across SRUS quantitative parameters and RECIST measurements, or a significant increase or no change. There was one exception: a participant showed a significant increase in vessel diameter and a significant decrease in blood flow speed and RECIST measurement at 6-months post-RT.

Fractal dimension, distance to vessel and distance to perimeter were also evaluated but fewer participants exhibited significant changes at 2-weeks and 6-months post-RT (Supplementary Figure 3).

### Correlation Between Post-Radiotherapy Changes of CD31 Vessel Count and SRUS Vessel Density

Overall, there was a reduction in blood vessels at 2-weeks post-RT as evidenced by the histopathological staining (Figure 8a) and corresponding SRUS microvascular map (Figure 8b) from the same participant. Additionally, the histopathological staining showed less conspicuous lumens, which corresponds to a reduction in SRUS vessel diameter.

**Figure 8.**
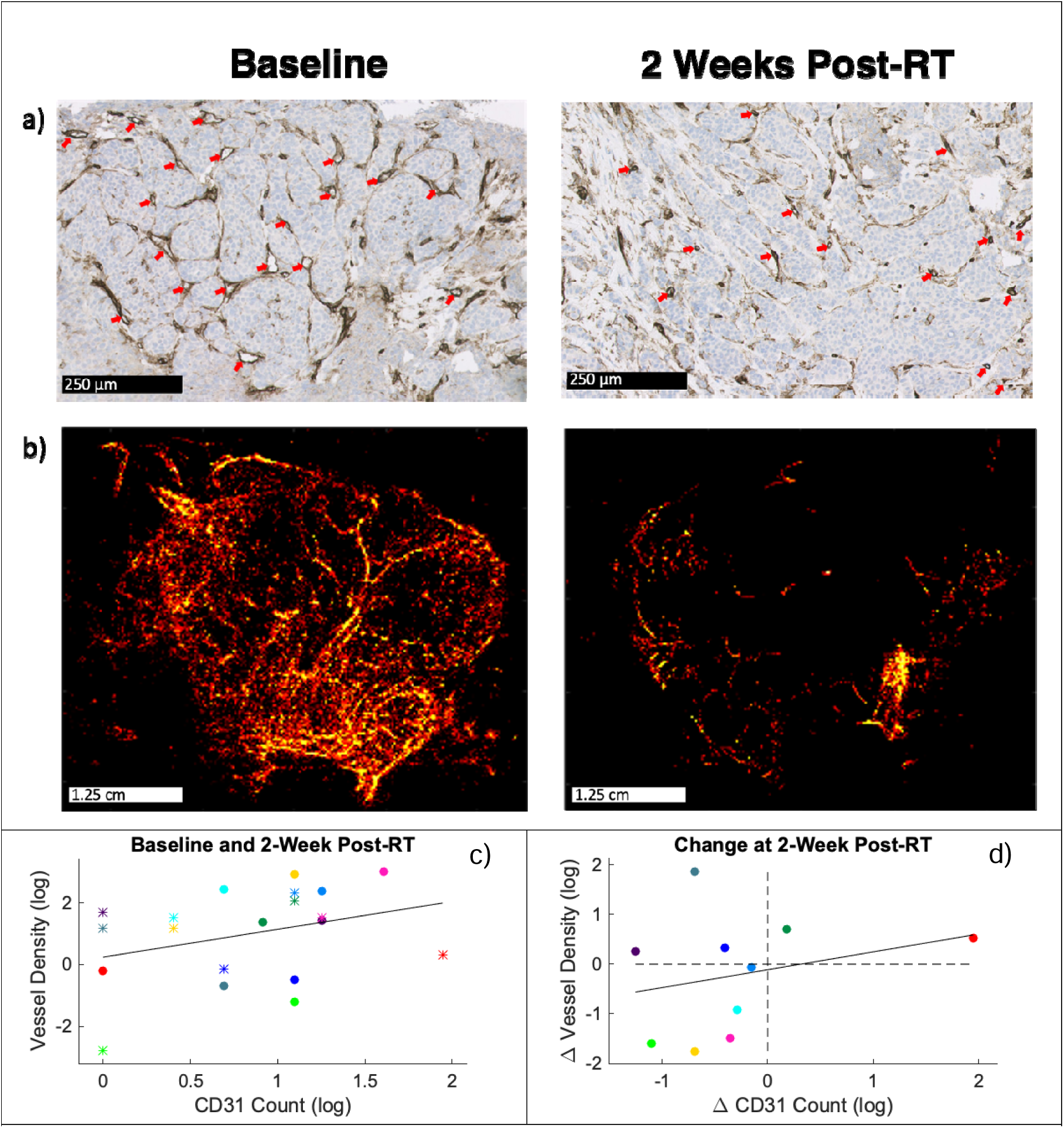
**a)** Breast core tumour biopsy section from a participant stained for CD31 (endothelial cell marker) at baseline (left) and 2-weeks post radiotherapy (right) showing a clear reduction in blood vessels. Some blood vessel examples are highlighted with red arrows; the stained vessels lumens generally appear smaller post-radiotherapy treatment. **b**) SRUS images from the same participant at baseline (left) and 2-weeks post-radiotherapy (right). **c**) Plot of the absolute values of the natural logarithm of the SRUS vessel density and CD31 count (as scored by a histopathologist), for 10 participants pre-radiotherapy (●) and 2-weeks post-radiotherapy (*), with the black line being a line of best fit. d) Plot of the change in the SRUS vessel density and CD31 count at 2-weeks post-RT, for 10 participants. The black line is the line of best fit, and the dashed black lines represent zero change. The colours signify the participants (matched with Figure 7).

Quantifying the immunohistochemical staining revealed a decrease in CD31 vessel count at 2-weeks post-RT for 8/10 participants that had core biopsies, as depicted in Figure 8d. Of these, 5 participants had a corresponding reduction in SRUS vessel density. Conversely, 2/10 participants showed an increase in CD31 vessel count at 2-weeks post-RT, with a corresponding increase in SRUS vessel density.

Visually, the absolute logarithmic values and changes in SRUS vessel density and CD31 vessel count showed a positive correlation (Figure 8c). However, due to the small sample size, statistical analysis was not performed. The plot for the absolute values includes data from two time-points for the same participants, differentiated by different markers (time-point) and colours (participant; matches colours in Figure 7).

## Discussion

The exceptional resolution and detail provided by super-resolution ultrasound (SRUS) offers significant potential for non-invasive detection of therapy-induced changes. Establishing SRUS’s repeatability and sensitivity to post-treatment changes is crucial for its clinical translation. Our study identified repeatable biomarkers of tumour microvasculature that were sensitive to early biological changes following completion of radiotherapy (RT). Notably, 5 of 11 participants showed significant SRUS biomarker alterations at 2-weeks post-RT. In comparison, only 1 of 11 participants had significant changes according to RECIST criteria, and this participant did not overlap with those showing SRUS changes. All 11 participants had a significant change by 2-weeks or 6-months post-RT in one or more SRUS quantitative parameters, compared to 6/11 participants with the RECIST measurement.

To our knowledge, this is the first evaluation of SRUS-derived biomarkers in response to radiotherapy. Acoustic angiography, a contrast-enhanced, high-resolution ultrasound microvascular imaging modality, has detected vascular changes earlier than changes in tumour size during the onset of recurrence in pre-clinical models (Kasoji 2018). Similarly, other imaging techniques, such as optoacoustic imaging (Wu 2022) and optical coherence tomography (Demidov 2018), have observed reductions in microvessel density following tumour irradiation, and subsequent revascularisation during recurrence, before any changes in tumour size. A microvasculature model revealed radiation-induced contraction of microvessels (Choi 2023). Additionally, in vivo micro-endoscopy (Byun 2019) detected significant decreases in blood flow speed following radiotherapy. Although these techniques validate our clinical findings, they are limited by shallow penetration depth and are predominately used in pre-clinical studies. Our research demonstrated that SRUS can similarly detect early vascular changes in patients, before any noticeable changes in tumour size. These promising findings support the potential of SRUS for long-term monitoring and early detection of treatment response, resistance and tumour recurrence. The destruction of the disordered microvasculature aligns with decreased local flow direction entropy; however, this entropy metric was less sensitive than other parameters in detecting response. Other SRUS-derived quantitative parameters (Opacic 2018, Zhu 2022), such as distance to vessel and fractal dimension, were initially investigated as potential indicators of radiotherapy response. However, these parameters showed minimal significant changes at 2-weeks and 6-months post-RT among the participants, indicating they were not effective predictors (included in Supplementary materials).

The heterogeneous microenvironment of solid tumours makes a single imaging plane insufficient to represent the entire tumour. Additionally, as tumour morphology changes post-therapy, it becomes challenging to return to the same imaging plane. This further emphasises the need for multiple imaging planes to adequately sample the tumour and obtain representative biomarkers for response monitoring. Our study demonstrated that averaging the SRUS quantitative parameters across multiple imaging planes improved their repeatability by reducing variations, thus yielding robust and reliable parameters. However, the number of imaging planes that can be acquired is limited by the decreasing microbubble concentration over time following the administration of a contrast bolus. In this study, 4 imaging planes were acquired within 2 minutes post-contrast administration, which is a feasible imaging timeframe in clinical practice.

To the best of our knowledge, no previous studies have examined SRUS repeatability in breast cancer. Related research on dynamic contrast-enhanced ultrasound imaging (Kaffas 2017, Feingold 2010) has shown that perfusion parameters from a single plane lack repeatability, and incorporating more planes improves it. The resolution of such images were diffraction-limited, so bulk tissue was assessed rather than individual micro-vessels.

For comparison, a contrast-enhanced ultrasound (CEUS) quantitative parameter was extracted, specifically, the sum of intensities within the maximum intensity projection. This CEUS parameter showed significant changes at 2-weeks and 6-months post-RT. This outcome was expected, as the parameter captures changes in microvascular density. However, CEUS alone misses vital functional information such as blood flow speed and direction. On the other hand, SRUS-derived blood flow speed proved important in evaluating response, with more participants showing significant changes than in SRUS-derived vessel density. The strength of SRUS lies in the abundance of information it provides on vascular structure and flow dynamics in superior resolution, which justifies its clinical evaluation with advantages over other imaging modalities.

SRUS-derived vessel density and CD31 vessel count from immunohistochemical staining showed a positive correlation, though statistical analysis was not conducted due to a small sample size. At 2-weeks post-RT, both measurements agreed in their direction of change for 7 out of 10 participants. Discrepancies may have arisen due to differences in sampling area: immunohistochemical staining sections cover millimetre-scale dimensions, while SRUS provides a wider field of view with centimetre-scale dimensions. By also imaging multiple planes, SRUS captures the heterogeneity of large breast tumours. Additionally, CD31 staining does not distinguish between viable and non-functional blood vessels, whereas SRUS visualises only functional vessels. However, some viable blood vessels might be undetected by SRUS. Solid tumours can experience acute or cyclic hypoxia due to intermittent blood flow, which can occur over minutes, a time-scale relevant to our repeatability study. Although not assessed in this study, SRUS could potentially investigate such hypoxia. SRUS might also miss the smallest blood vessels during imaging because microbubbles are less likely to pass through them within the short acquisition time. Other biological variations, such as heart-rate, could affect microbubble concentration and blood flow speed, but these variations should be minimal in a resting position.

Biases in SRUS quantitative parameters can also arise from technical factors. Hardware constraints led to a trade-off between compounded frame-rate, acquisition length, and the number of compounded angles, due to data collection limits per acquisition. While longer acquisitions can improve parameter robustness by increasing vascular saturation, they also pose challenges such as greater motion artifacts, necessitating customised setups to fix the probe. A fixed probe, though, limits the flexibility needed to quickly locate the tumour’s central slice when moving between imaging planes, and does not mitigate patient motion. Moreover, extended acquisition times mean that later imaging planes are captured further from the concentration peak. Higher frame-rates improve tracking capabilities for estimating blood flow speed and direction, and future work will investigate the impact of frame-rate on SRUS quantitative parameters. Nevertheless, our study’s acquisition time and frame-rate were sufficient for detecting post-therapy changes in SRUS quantitative parameters. Microbubble tracking accuracy can influence blood flow speed and flow direction parameters, where accuracy may be reduced in denser microvasculature or fast-flowing blood vessels. Further development and benchmarking in SRUS localisation and tracking algorithms, which was the aim of a recent international challenge (Lerendegui 2022), will increase quantitative parameter robustness.

Statistical limitations included using the same participant at multiple time-points. The repeatability scans from the same participants were considered as independent data-points during statistical analysis, due to the length of time between repeatability scans and post-treatment changes occurring over that time. Typically, repeatability studies for medical imaging technologies involve conducting repeat scans on different days. However, this study was limited to a single imaging session, due to scheduling issues, including the need to start radiotherapy treatment soon after the baseline scan. Before administering the second bolus, microbubble concentration must diminish to negligible levels. Any residual circulating microbubbles could increase the concentration of the second bolus, and impact concentration-sensitive SRUS quantitative parameters. Before injecting the second bolus, the clearance of the first bolus was visually confirmed on the real-time feedback. A 5 minute-interval between bolus injections allowed for the microbubbles to clear.

The repeatability coefficient calculated in this study reflects the technical variability of SRUS and its influence on SRUS-derived quantitative parameters. This technical variability remained consistent both pre-treatment and at various post-treatment time-points. Additionally, biological variability occurring over a 5-minute interval also contributes to the repeatability coefficient magnitude. The changes observed at later time-points in Figure 7 exceed the variability seen when measurements are taken five minutes apart. However, we suggest interpreting Figure 7 with caution. While it shows the changes in some metrics before and after RT is larger than the variations when measurements are taken five minutes apart, it is possible that a measurement taken with longer intervals, even when the tumour is unchanged in the meantime, will have a different distribution of variations. This is a preliminary study and any conclusion of significant changes after RT is dependent on long term variation in measurement being the same as the very short-term differences that have been demonstrated, something that needs future verification.

Despite the typical practice of pre-treatment repeatability studies (O’Connor 2017), we wanted to also ensure SRUS repeatability at post-treatment timepoints, and we believe this approach strengthens our study. From the Bland-Altman plots, there was no clear evidence that repeatability varied across imaging time-points, however, repeatability was heteroskedastic, i.e., the variability was related to the mean, with more vascular tumours exhibiting relatively poorer absolute repeatability. Post-treatment repeatability studies have also been conducted using MRI (Newitt 2019), lending further support to our study design.

Contrast-enhanced ultrasound with SonoVue is clinically licensed for diagnosing liver metastases, coronary artery disease and vesicoureteral reflux among others, and is a cost-effective, safe, bedside imaging modality. SRUS inherits these advantages and holds the potential for seamless clinical translation. A standardised protocol and user-friendly software are essential for SRUS to be employed in therapy response monitoring. While validated here in breast tumours, SRUS repeatability must be established for other tumours, and its potential in clinically monitoring microvasculature changes to other anti-cancer therapies, which has been investigated pre-clinically (Hoyt 2024, Yin 2024, Brown 2023, Lowerison 2022). This study only explored intra-operator variability; investigating inter-operator variability and conducting multi-centre studies will further establish the repeatability and reproducibility of SRUS.

In summary, this first-in-patient repeatability study has demonstrated that SRUS provides robust biomarkers quantifying tumour microvasculature structure and blood flow dynamics when assessing short-term, technical variability. These highly repeatable SRUS-derived quantitative parameters are sensitive to early biological changes following treatment, with SRUS vessel density validated against gold-standard histopathological vessel count scores. To confirm if changes in the SRUS-derived quantitative parameters are early indicators of radiotherapy response, correlation with long-term radiological response is required. The results of this preliminary study are encouraging, showcasing SRUS’s potential as an affordable non-invasive monitoring tool in identifying response, resistance, and relapse earlier than is currently achievable.

## Data Availability

All data produced in the present study are available upon reasonable request to the authors.

## Contributors

Conceptualisation: MM, ED, SA, KD, MDB, NS and MXT. Formal analysis: MM, ED and IR. Funding acquisition: MDB, NS and MXT. Investigation: MM, ED, VS, IR, JS, TG, PSM, SN, JHS and NS. Project administration: LG and JH. Software: MM, MT, JY and BH. Supervision: MDB, NS and MXT. Visualisation: MM and ED. Writing – original draft: MM and ED. Writing – review & editing: all authors. All authors have read and approved the final version of the manuscript.

## Data Sharing Statement

Data will be made available upon reasonable request to Mengxing Tang and Navita Somaiah.

## Acknowledgements

This research project and MM is supported by the CRUK Convergence Science Centre at The Institute of Cancer Research, London, and Imperial College London (A26234). MT is funded by NIHR i4i under Grant No. NIHR200972, BH is funded by China Scholarship Council, and JY is funded by Chan Zuckerberg Foundation under Grant No. 2020-225443. We acknowledge funding from Kortuc Inc., Japan, NHS funding to the NIHR Biomedical Research Centre at The Royal Marsden NHS Foundation Trust and the Institute of Cancer Research, the National Institute for Health Research (grant No. NIHR200972) and the ICiC & IAA funding from Imperial College. Thank you to the healthcare staff at the Rapid Diagnostic Centres at the Royal Marsden, and the patients who kindly agreed to participate in this study.

