## Supplementary Material for "In-Patient Repeatability and Sensitivity Study of Multi-Plane Super-Resolution Ultrasound in Breast Cancer"

**Supplementary methods**

***Quantitative Parameters***

**Fractal Dimension.** Fractal dimension was calculated using a box-counting method (available online: (<https://www.mathworks.com/matlabcentral/fileexchange/13063-boxcount>). This method determines the number of boxes of varying sizes required to cover the entire microvascular structure. The fractal dimension is then obtained as the slope of the log-log plot of the box size against the number of boxes needed to cover the structure for each box size. The fractal dimension measures structural complexity; tumour microvasculature is chaotic in comparison to the ordered blood vessels in healthy tissue.

**Distance to Vessel.** The distance from each pixel within the delineated tumour, excluding those containing blood vessels, to the nearest blood vessel edge (either inside or outside the tumour) was calculated (bwdist, MATLAB, Image Processing Toolbox). The in-plane mean was then determined by averaging these values across the tumour. The tumour microvasculature is chaotic, characterised by areas of dense blood vessel distribution.

**Distance to Perimeter.** The normalised distance from each blood vessel pixel within the tumour to the tumour perimeter, relative to the tumour area was calculated. The perimeter was extracted (bwperim, MATLAB, Image Processing Toolbox), and the distance from blood vessels to the perimeter was determined (bwdist, MATLAB, Image Processing Toolbox). Tumours typically exhibit dense microvasculature at the periphery, while the core is often poorly perfused.

**Supplementary results**

| 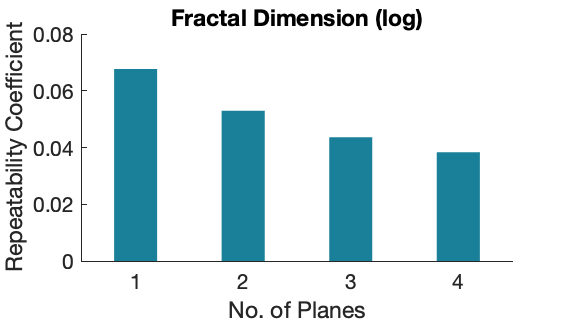  -43%  -36%  -22% | 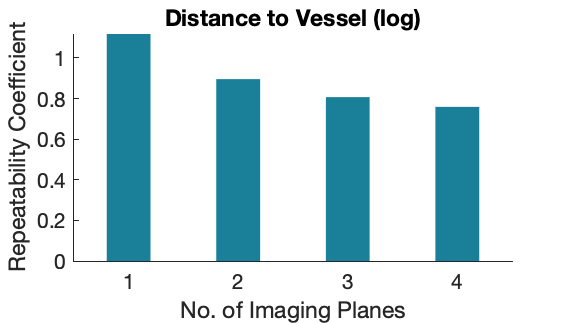  -32%  -28%  -20% |
| --- | --- |
| 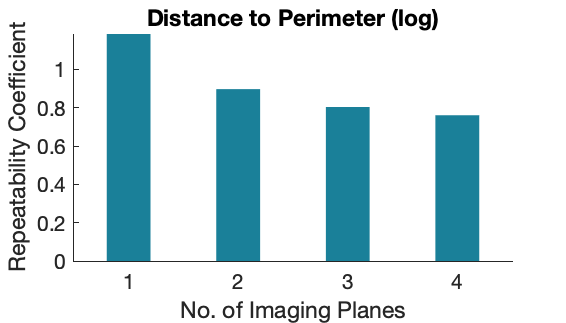  -36%  -32%  -24% | |
| **Supplementary Figure 1.** The repeatability coefficient (RC) of the natural logarithm of SRUS quantitative parameters, as a function of the number of imaging planes used for the average measure. The percentages displayed above the bars represent the percentage change compared to using 1 imaging plane. | |

| 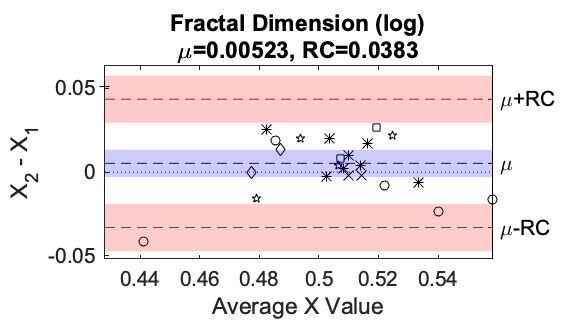 | 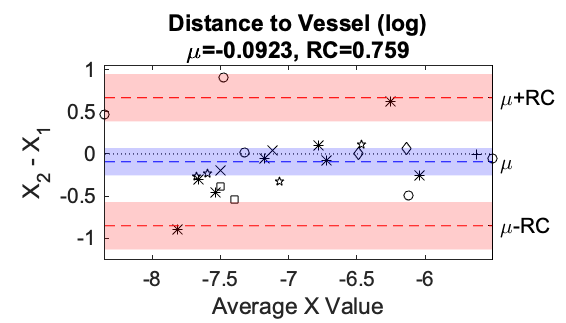 |
| --- | --- |
| 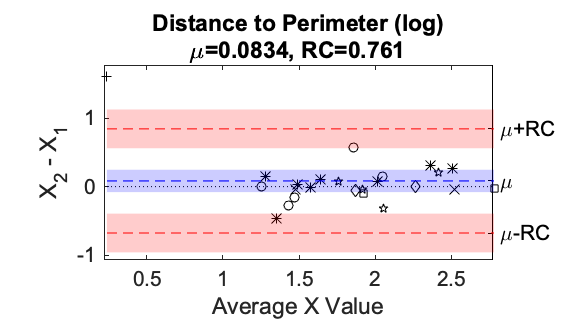 | |
| **Supplementary Figure 2.** Bland-Altman plots for 24 repeatability scans. The natural logarithm of the SRUS quantitative parameters is used for statistical analysis. The y-axis is the difference between the quantitative parameter X, averaged across all 4 imaging planes, from the 2 repeat measurements (after the first bolus X_1_ and second bolus X_2_). The x-axis is the average value of the 2 repeat measurements of the quantitative parameter. The mean of the differences ($\mu$) is given as the blue dashed line, with its 95% confidence interval being the shaded blue area. The dotted line represents 0 difference between the test-retest measurement. The mean of the differences +/- the repeatability coefficient ($\mu\pm RC$) is given as the red dashed line, with the 95% confidence interval of the repeatability coefficient represented by the shaded red area. The markers are the individual repeatability scans (N=24), with ○ representing baseline scans; * 2-weeks post-RT; + 3-months post-RT; ☆ 6-months post-RT; × 12-months post-RT; □ 18-months post-RT; and ◇ 24-months post-RT. | |

| 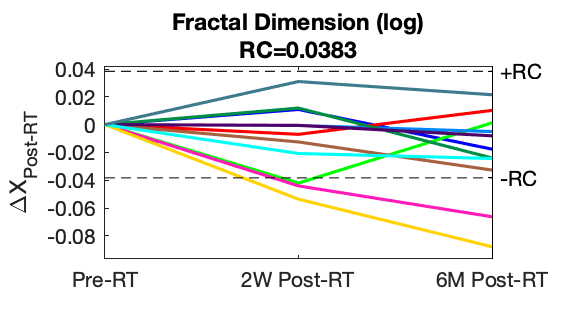 | 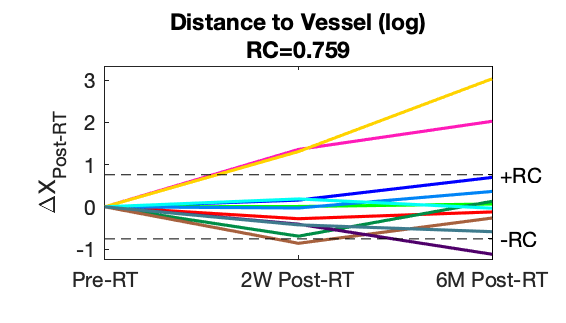 |
| --- | --- |
| 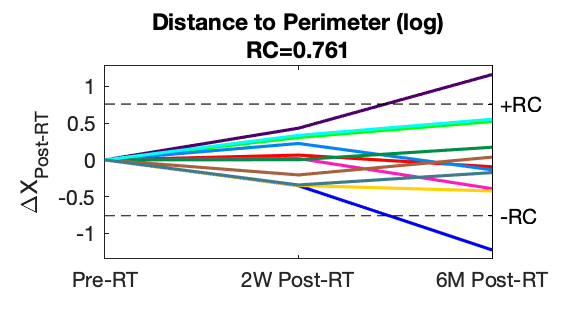 | |
| **Supplementary Figure 3.** a) The changes of the natural logarithm of the super-resolution ultrasound (SRUS) quantitative parameters 2-weeks and 6-months post-radiotherapy (RT) from 11 participants. The y-axis shows the change in the mean of the quantitative parameter (averaged across 4 imaging planes) 2-weeks and 6-months post-RT from baseline. The dashed black lines represent the repeatability coefficient, as indicator of significant change, i.e., if the difference post-treatment is larger than the repeatability coefficient, then the change is significant. | |
